# Association of Bacillus Calmette-Guerin (BCG) vaccination with reduced risk of PFAPA: nationwide matched case-control study using electronic health records

**DOI:** 10.64898/2026.01.04.26343402

**Authors:** Ariel Israel, Ilan Green, Shai Ashkenazi, Eugene Merzon, Avivit Golan-Cohen, Shlomo Vinker, Eli Magen, Yackov Berkun

## Abstract

**Objectives:** Periodic fever, aphthous stomatitis, pharyngitis, and cervical adenitis (PFAPA) syndrome is the most common autoinflammatory disorder of childhood, characterized by recurrent febrile episodes driven by dysregulated innate immune activation. Although genetic susceptibility contributes to disease risk, environmental modifiers remain poorly defined. Vaccinations may induce long-lasting modulation of innate immune responses and influence PFAPA incidence. We aimed to assess whether childhood vaccinations, including Bacillus Calmette-Guerin (BCG), are associated with PFAPA risk in a large national cohort.

**Methods:** We conducted a matched case-control study using electronic health records from a national healthcare provider in Israel. Children diagnosed with PFAPA were matched to children without PFAPA by age, sex, socioeconomic status, ethnic sector, and enrolment year. Vaccination history was obtained from the electronic immunization registry. A systematic screen of associations was performed across all recorded childhood vaccines, followed by adjusted conditional logistic regression models. Prior laboratory test results were analysed to characterize the immune profile.

**Results:** The study included 1,641 PFAPA cases and 32,820 matched controls. Among all vaccines examined, prior Bacillus Calmette-Guerin (BCG) vaccination, received by 401 children (3 cases, 398 controls), showed the strongest association with reduced PFAPA risk (adjusted odds ratio 0.15, 95% confidence interval [CI] 0.05-0.46, p=0.001). Associations with other vaccines were heterogeneous and of smaller magnitude. PFAPA cases demonstrated a myeloid-skewed inflammatory profile prior to diagnosis, including elevated C-reactive protein, neutrophilia, increased neutrophil-to-lymphocyte ratio, and relative lymphopenia.

**Conclusions:** In this nationwide study, prior BCG vaccination emerged from a systematic screen as strongly associated with reduced PFAPA risk, supporting a role for immune programming in susceptibility to childhood autoinflammatory disease.

## Introduction

Periodic fever, aphthous stomatitis, pharyngitis, and cervical adenitis (PFAPA) syndrome is the most common autoinflammatory disorder of childhood. It is characterized by stereotyped episodes of high fever accompanied by mucosal and lymphoid inflammation, with complete clinical remission between attacks. Despite being described more than three decades ago, the pathogenesis of PFAPA remains incompletely understood.[1–5] In routine paediatric practice, children with PFAPA frequently undergo repeated assessments for suspected infection and may receive multiple courses of empirical antibiotics before the diagnosis is recognised. Understanding potentially modifiable determinants of PFAPA risk is therefore relevant to paediatric infectious disease management and antibiotic stewardship.

Accumulating evidence supports a central role for dysregulated innate immune activation in PFAPA. Febrile episodes are associated with increased production of proinflammatory cytokines, including interleukin-1beta and interleukin-18, consistent with inflammasome activation and downstream Th1-skewed immune responses. Tonsillar tissue is thought to serve as a key site of immune triggering and cytokine amplification, supported by the clinical efficacy of tonsillectomy in a substantial proportion of patients.[5–9]

Although genetic susceptibility contributes to PFAPA risk, the disorder is not monogenic, suggesting an important role for environmental modifiers. Early-life exposures that shape innate immune development during critical windows may influence later susceptibility to autoinflammatory flares[8,9]. Among such exposures, Bacillus Calmette-Guerin (BCG) vaccination is of particular interest. Beyond its role in tuberculosis prevention, BCG induces durable functional reprogramming of innate immune cells through epigenetic and metabolic mechanisms, a phenomenon commonly referred to as trained immunity.[10–12]

Evaluating the long-term immunologic associations of BCG vaccination in contemporary Western populations is increasingly challenging, as routine BCG administration has been discontinued in most countries where large-scale electronic health record (EHR) systems are available. However, in healthcare systems where BCG vaccination remains selectively administered to defined risk groups, such as Israel, observational EHR-based studies provide a distinctive opportunity to examine long-term immunologic associations of BCG in non-endemic, high-income settings.

Using comprehensive EHR data from a nationwide healthcare organization in Leumit Health Services (LHS), a national healthcare provider in Israel, we took advantage of this unique setting to assess associations between vaccinations, including early-life BCG vaccination, and PFAPA risk. Systematically recorded vaccination histories, including documentation imported at enrolment for individuals vaccinated in countries with ongoing BCG programs, enabled evaluation of this association in a non-tuberculosis-endemic setting. We further contextualized these findings using laboratory markers of immune activation preceding PFAPA diagnosis.

## Methods

### Study Design and Data Source

We conducted a nationwide, retrospective, matched case-control study using electronic health records (EHRs) from LHS, one of Israel’s four national health maintenance organizations, providing care to approximately 730,000 members. LHS maintains a centralized longitudinal EHR system spanning more than two decades, including demographics, diagnoses, vaccinations, laboratory tests, medication prescriptions and purchases, and healthcare utilization. All Israeli residents are covered under a universal healthcare system with standardized benefits and a national immunization program. Diagnostic coding in LHS is entered by treating physicians using ICD-9 codes, has been validated in multiple peer-reviewed studies, and allows diagnoses to be designated as chronic when they reflect ongoing conditions.[13,14] Chronic diagnoses undergo active longitudinal curation, and treating physicians may subsequently remove diagnoses determined to be incorrect.

### Study Population and Case Definition

The source population included all LHS members born between 2000 and 2024. PFAPA cases were identified based on the presence of a uniquely defined PFAPA diagnosis code (ICD-9 code 277.38), a locally implemented code available for selection in the EHR and used by LHS physicians to document PFAPA as a chronic diagnosis. To enhance diagnostic specificity, we included only chronic diagnoses recorded between ages 1 and 9 years that remained active and had not been subsequently removed from the medical record. The date of the first recorded PFAPA diagnosis served as the index date. Individuals with incomplete enrolment history before the index date were excluded. Sex was recorded in the electronic health record as male or female, as assigned at birth

### Control Selection, Matching, and Calendar Alignment

For each PFAPA case, 20 controls without a PFAPA diagnosis were selected using exact matching on sex, birth year, ethnic sector, socioeconomic status, and year of first recorded healthcare contact. Controls were required to be alive and actively enrolled in LHS at the case’s index date. To ensure strict calendar alignment and avoid immortal-time or temporal bias, the index date of each case was assigned to its matched controls, and all exposures and laboratory measurements were ascertained only from time windows preceding the assigned index date. Cases for which sufficient exact matches could not be identified were excluded. The final analytical cohort included 1,641 PFAPA cases and 32,820 matched controls. Cohort construction, inclusion criteria, and exclusions are detailed in **Figure 1**, which demonstrates temporal alignment and minimizes selection and temporal bias.

**Figure 1.**
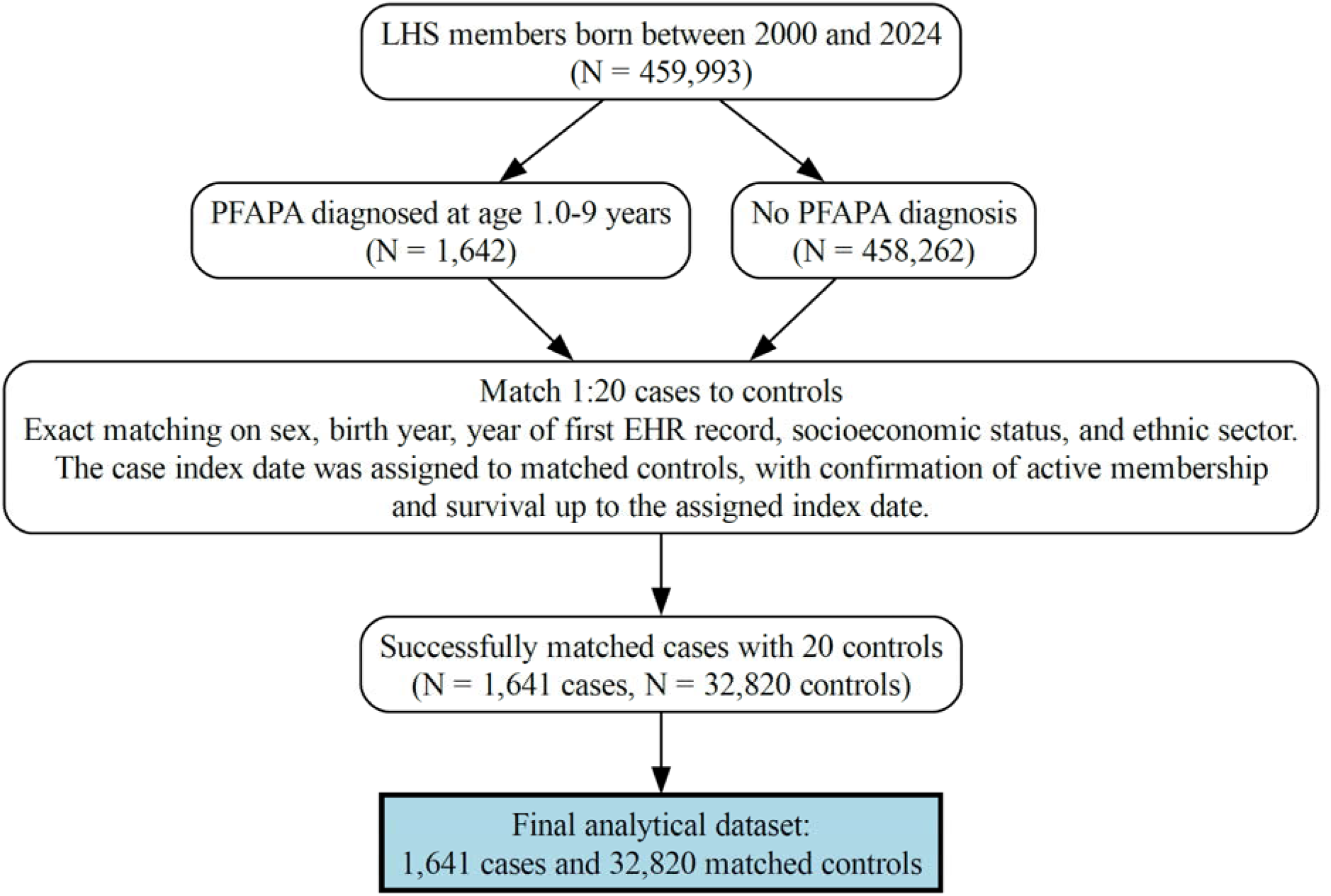
Cohort construction flowchart. Among Leumit Health Services (LHS) members born between 2000 and 2024 (N = 459,993), PFAPA cases were defined as individuals diagnosed between ages 1 and 9 years (N = 1,642) and were matched 1:20 to controls without a PFAPA diagnosis (N = 458,262). Exact matching was performed on sex, birth year, year of first EHR record, socioeconomic status, and ethnic sector. Each control was assigned the index date of the matched case, with confirmation of active LHS membership and survival through the assigned index date. Of 1,642 PFAPA cases identified, 1,641 were successfully matched and included in the analysis. The final analytical dataset included 1,641 PFAPA cases and 32,820 matched controls.

### Vaccination Exposure Assessment

Vaccination history was extracted from the LHS immunization registry. In Israel, Bacillus Calmette-Guerin (BCG) vaccination is not part of the universal childhood immunization schedule and is administered selectively according to Ministry of Health guidelines. During the study period, BCG vaccination was recommended for newborns and children up to age 4 years who met predefined risk criteria, primarily related to the country of origin of the child or parents, prior residence in high tuberculosis-incidence settings, or household exposure to tuberculosis. Eligible newborns born in Israel are typically vaccinated in hospital shortly after birth, whereas catch-up vaccination may be administered in community well-child clinics for eligible infants or children, including those born abroad. Vaccinations administered outside Israel may also be documented retrospectively in the immunization registry upon enrolment in the health system.

All routinely recorded childhood vaccines were evaluated. An unsupervised screen of associations between prior vaccination status and PFAPA was performed, without prespecifying a primary vaccine exposure.

### Laboratory Markers

Laboratory test results obtained before the index date were analysed to characterize immune and inflammatory profiles. For each laboratory marker, predefined summary measures (e.g., maximum, minimum, or last recorded value) were extracted within the pre-diagnostic window, as specified a priori, to avoid influence of diagnostic workup or treatment initiation.

### Statistical Analysis

Baseline characteristics were summarized using means with standard deviations or medians with interquartile ranges (IQR), as appropriate, and counts with percentages for categorical variables. Covariate balance between cases and controls was assessed using standardized mean differences (SMDs), which are independent of sample size and statistical significance testing; p-values for baseline comparisons are therefore reported for descriptive purposes only.

An exploratory screening step was performed using Fisher’s exact test to compare vaccination coverage between cases and controls across 35 childhood vaccines recorded in the EHR. False discovery rate correction using the Benjamini–Hochberg procedure was applied to the unadjusted screening results. Vaccines meeting a prespecified unadjusted screening threshold of P < 0.10 were selected for further evaluation. For these vaccines, a multivariable conditional logistic regression model was used to estimate adjusted odds ratios (aORs) and 95% confidence intervals (CIs), accounting for the matched design. The model simultaneously included all screened vaccines, allowing estimation of vaccine-specific associations while accounting for correlated vaccination patterns and overlapping indications that could otherwise act as confounders. The model additionally adjusted for residual variation in age, sex, ethnic sector, and socioeconomic status arising from categorization and within-stratum heterogeneity.

All p-values are two-sided, with P < 0.05 considered statistically significant. Analyses were conducted using R version 4.4.0, with data extraction and preprocessing performed using structured query language (SQL) and Python version 3.11.

A large language model (ChatGPT 5) was used for language editing and clarity; all authors reviewed and take responsibility for the final content.

### Ethics and Reporting

In accordance with the Declaration of Helsinki, the study protocol was approved by the Leumit Health Services Institutional Review Board (LEU-23-11), which granted a waiver of informed consent because analyses were conducted on de-identified data. Reporting followed the STROBE guideline and its RECORD extension for studies using routinely collected health data.

## Results

### Cohort assembly

**Figure 1** presents the cohort construction flowchart. Among 459,993 individuals born between 2000 and 2024 and insured by LHS, 1,641 children diagnosed with PFAPA between ages 1 and 9 years were identified and successfully matched to 32,820 controls without PFAPA using exact matching criteria.

### Baseline characteristics

Baseline demographic and anthropometric characteristics are summarized in Table 1. Matching variables were identical by design, and anthropometric measures were similar between groups.

**Table 1.**
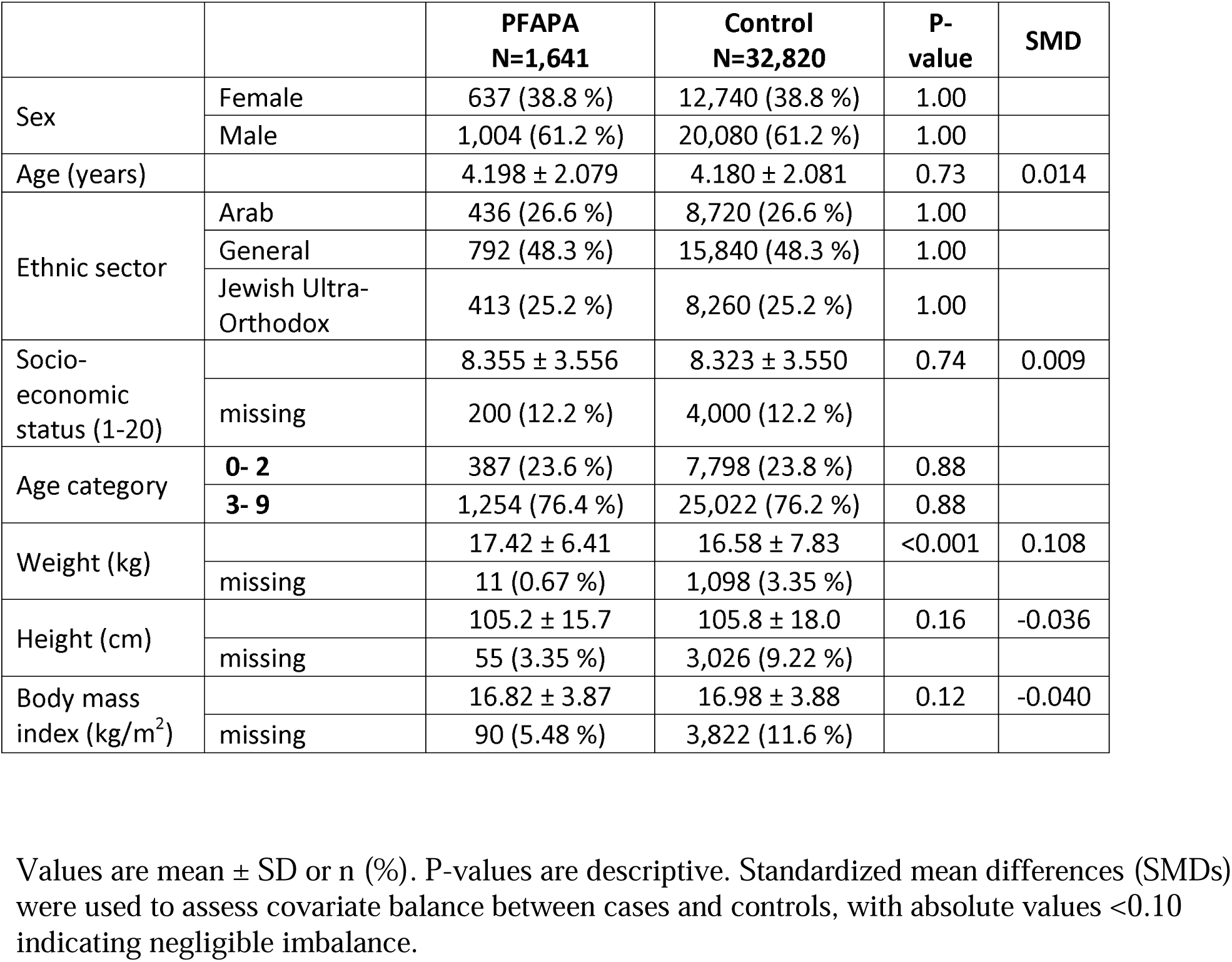
Demographic and clinical characteristics of the study cohort at index date.

### Pre-diagnostic inflammatory laboratory profile

Selected laboratory markers obtained within 30 days preceding diagnosis are presented in **Table 2**. PFAPA cases demonstrated a consistent inflammatory and myeloid-skewed profile. These findings support the presence of innate immune activation near the time of diagnosis.

**Table 2.**
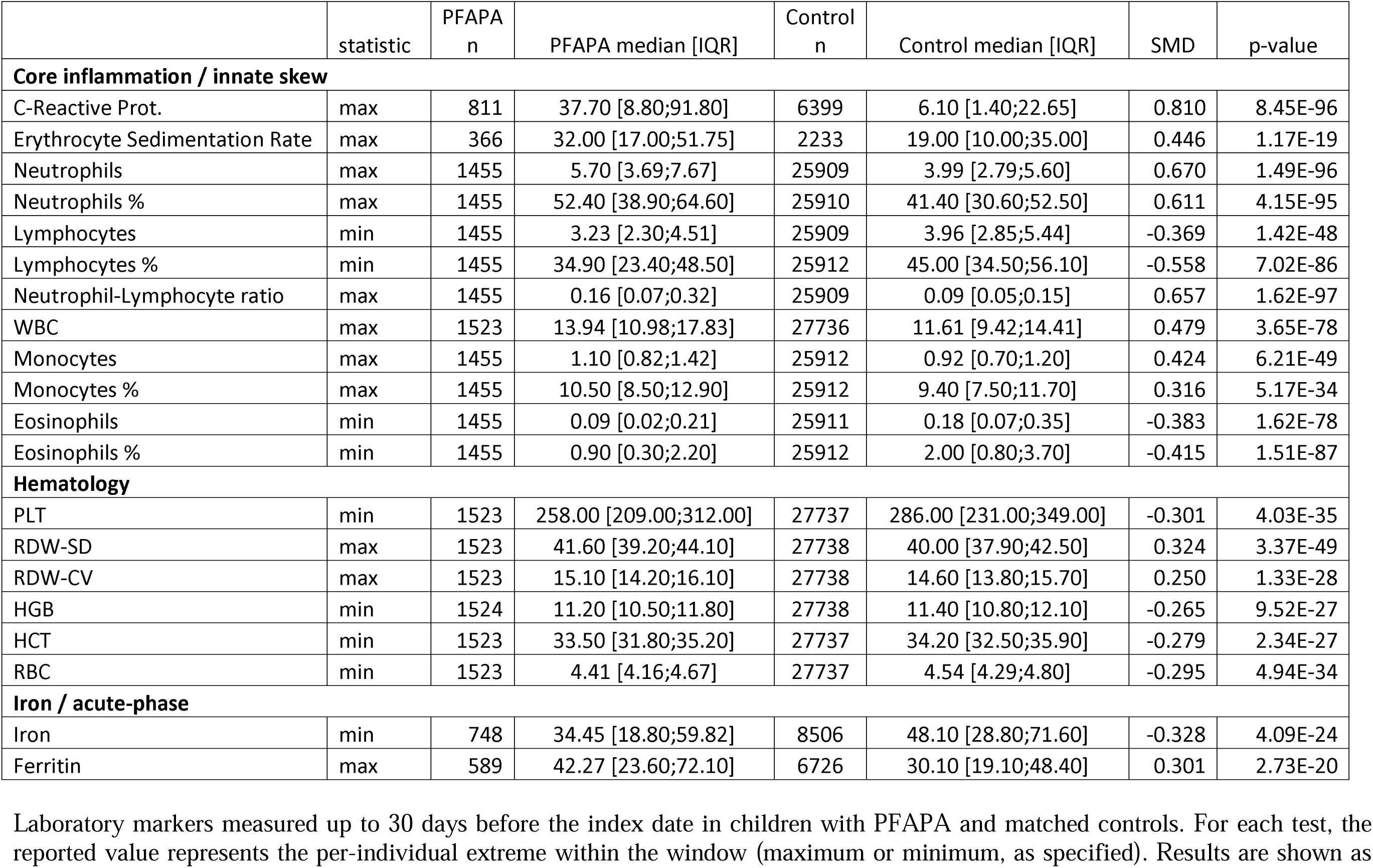
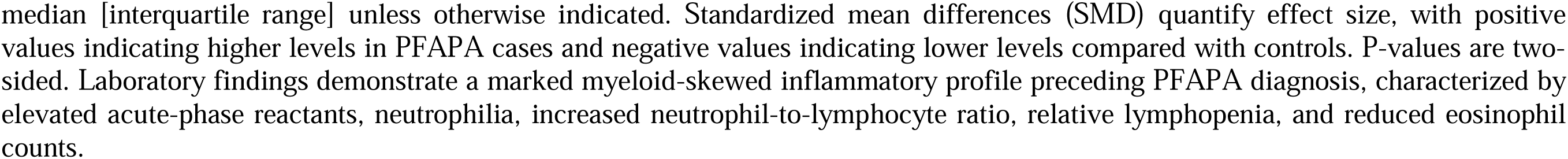
Prediagnostic laboratory features associated with PFAPA.

|  | statistic | PFAPA<br>n | PFAPA median [IQR] | Control<br>n | Control median [IQR] | SMD | p-value |
| --- | --- | --- | --- | --- | --- | --- | --- |
| **Core inflammation / innate skew** |  |  |  |  |  |  |  |
| C-Reactive Prot. | max | 811 | 37.70 [8.80;91.80] | 6399 | 6.10 [1.40;22.65] | 0.810 | 8.45E-96 |
| Erythrocyte Sedimentation Rate | max | 366 | 32.00 [17.00;51.75] | 2233 | 19.00 [10.00;35.00] | 0.446 | 1.17E-19 |
| Neutrophils | max | 1455 | 5.70 [3.69;7.67] | 25909 | 3.99 [2.79;5.60] | 0.670 | 1.49E-96 |
| Neutrophils % | max | 1455 | 52.40 [38.90;64.60] | 25910 | 41.40 [30.60;52.50] | 0.611 | 4.15E-95 |
| Lymphocytes | min | 1455 | 3.23 [2.30;4.51] | 25909 | 3.96 [2.85;5.44] | -0.369 | 1.42E-48 |
| Lymphocytes % | min | 1455 | 34.90 [23.40;48.50] | 25912 | 45.00 [34.50;56.10] | -0.558 | 7.02E-86 |
| Neutrophil-Lymphocyte ratio | max | 1455 | 0.16 [0.07;0.32] | 25909 | 0.09 [0.05;0.15] | 0.657 | 1.62E-97 |
| WBC | max | 1523 | 13.94 [10.98;17.83] | 27736 | 11.61 [9.42;14.41] | 0.479 | 3.65E-78 |
| Monocytes | max | 1455 | 1.10 [0.82;1.42] | 25912 | 0.92 [0.70;1.20] | 0.424 | 6.21E-49 |
| Monocytes % | max | 1455 | 10.50 [8.50;12.90] | 25912 | 9.40 [7.50;11.70] | 0.316 | 5.17E-34 |
| Eosinophils | min | 1455 | 0.09 [0.02;0.21] | 25911 | 0.18 [0.07;0.35] | -0.383 | 1.62E-78 |
| Eosinophils % | min | 1455 | 0.90 [0.30;2.20] | 25912 | 2.00 [0.80;3.70] | -0.415 | 1.51E-87 |
| **Hematology** |  |  |  |  |  |  |  |
| PLT | min | 1523 | 258.00 [209.00;312.00] | 27737 | 286.00 [231.00;349.00] | -0.301 | 4.03E-35 |
| RDW-SD | max | 1523 | 41.60 [39.20;44.10] | 27738 | 40.00 [37.90;42.50] | 0.324 | 3.37E-49 |
| RDW-CV | max | 1523 | 15.10 [14.20;16.10] | 27738 | 14.60 [13.80;15.70] | 0.250 | 1.33E-28 |
| HGB | min | 1524 | 11.20 [10.50;11.80] | 27738 | 11.40 [10.80;12.10] | -0.265 | 9.52E-27 |
| HCT | min | 1523 | 33.50 [31.80;35.20] | 27737 | 34.20 [32.50;35.90] | -0.279 | 2.34E-27 |
| RBC | min | 1523 | 4.41 [4.16;4.67] | 27737 | 4.54 [4.29;4.80] | -0.295 | 4.94E-34 |
| **Iron / acute-phase** |  |  |  |  |  |  |  |
| Iron | min | 748 | 34.45 [18.80;59.82] | 8506 | 48.10 [28.80;71.60] | -0.328 | 4.09E-24 |
| Ferritin | max | 589 | 42.27 [23.60;72.10] | 6726 | 30.10 [19.10;48.40] | 0.301 | 2.73E-20 |

Laboratory markers measured up to 30 days before the index date in children with PFAPA and matched controls. For each test, the reported value represents the per-individual extreme within the window (maximum or minimum, as specified). Results are shown as
median [interquartile range] unless otherwise indicated. Standardized mean differences (SMD) quantify effect size, with positive values indicating higher levels in PFAPA cases and negative values indicating lower levels compared with controls. P-values are two-sided. Laboratory findings demonstrate a marked myeloid-skewed inflammatory profile preceding PFAPA diagnosis, characterized by elevated acute-phase reactants, neutrophilia, increased neutrophil-to-lymphocyte ratio, relative lymphopenia, and reduced eosinophil counts.

### Association between childhood vaccinations and PFAPA

An exploratory, unsupervised screening was performed to identify candidate vaccines for adjusted analysis (**Table 3**). Among all vaccines evaluated, BCG vaccination showed a markedly lower prevalence among PFAPA cases compared with controls and was the only vaccine to remain statistically significant after false discovery rate correction. It was therefore the primary focus of subsequent multivariable analyses.

**Table 3:** Unadjusted associations between childhood vaccinations and PFAPA.

| Injection | PFAPA<br>vaccinated<br>n (%) | Control<br>vaccinated<br>n (%) | Odds ratio | 95% CI | P-value |
| --- | --- | --- | --- | --- | --- |
| BCG | 3 (0.18) | 398 (1.21) | 0.15 | [0.03-0.44] | <0.00001 |
| ROTA | 996 (60.7) | 18,690 (56.9) | 1.17 | [1.05-1.29] | 0.003 |
| bOPV | 1,022 (62.3) | 19,517 (59.5) | 1.13 | [1.01-1.25] | 0.023 |
| OPV | 76 (4.6) | 1,452 (4.4) | 1.05 | [0.82-1.33] | 0.67 |
| DTaP-HIB+IPV | 1,389 (84.6) | 27,157 (82.7) | 1.15 | [1.00-1.32] | 0.048 |
| DTaP-HIB+IPV Poliacel | 20 (1.2) | 625 (1.9) | 0.64 | [0.38-0.99] | 0.049 |
| DTaP-HIB+IPV Pediacel | 481 (29.3) | 9,667 (29.5) | 0.99 | [0.89-1.11] | 0.91 |
| DTaP-HIB+IPV Pentaxim | 200 (12.2) | 3,997 (12.2) | 1.00 | [0.86-1.17] | 1.00 |
| MMR | 210 (12.8) | 4,405 (13.4) | 0.95 | [0.81-1.10] | 0.50 |
| MMRV | 1,182 (72.0) | 23,369 (71.2) | 1.04 | [0.93-1.17] | 0.49 |
| HAV | 1,249 (76.1) | 24,354 (74.2) | 1.11 | [0.99-1.25] | 0.09 |
| HBV | 1,356 (82.6) | 26,766 (81.6) | 1.08 | [0.94-1.23] | 0.28 |
| PCV13 | 1,112 (67.8) | 21,662 (66.0) | 1.08 | [0.97-1.21] | 0.15 |

Unadjusted associations between prior childhood vaccination and PFAPA in a matched case-control cohort. Odds ratios and 95% confidence intervals were calculated from contingency tables; p-values were obtained using Fisher's exact test. This table represents an unsupervised screening analysis performed across all routinely recorded childhood vaccines. No correction for multiple testing was applied. Inference is based on multivariable conditional logistic regression models presented separately. OPV = trivalent oral polio vaccine (historical) bOPV = bivalent oral polio vaccine (later formulation)

Vaccines meeting the prespecified screening threshold were subsequently evaluated in a multivariable conditional logistic regression model accounting for the matched design (**Figure 2**). In the adjusted model, BCG vaccination remained strongly associated with reduced PFAPA risk. In addition, MMR, MMRV, and the DTaP-HIB+IPV (Poliacel) formulation were associated with modestly lower odds of PFAPA, whereas rotavirus vaccination and bivalent oral polio vaccine showed higher odds of PFAPA.

**Figure 2.**
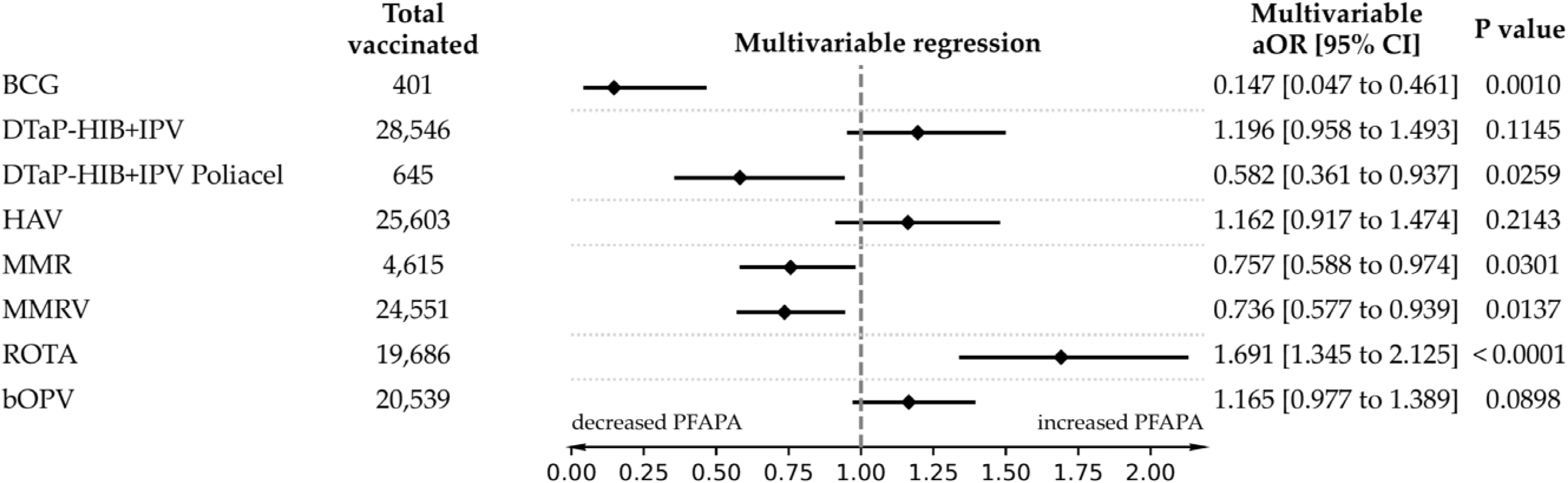
Adjusted associations between selected childhood vaccinations and PFAPA risk. Forest plot showing adjusted odds ratios (aORs) and 95% confidence intervals for the association between selected childhood vaccinations and PFAPA, estimated using conditional logistic regression. Vaccines shown passed an initial unadjusted screening threshold (P < 0.10) in Fisher’s exact tests. Models were adjusted for residual differences in age, sex, ethnic sector, and socioeconomic status within matched strata. Odds ratios <1 indicate reduced PFAPA risk.

## Discussion

In this large nationwide matched case-control study using comprehensive electronic health records, early-life BCG vaccination was strongly associated with a reduced risk of PFAPA. Among all routinely recorded childhood vaccines, BCG emerged as the strongest protective association in an unsupervised screening analysis and remained highly significant in multivariable conditional logistic regression. This association was observed in a setting where BCG is not administered routinely, but selectively to individuals meeting predefined risk criteria, primarily related to the country of origin of the child or parents or prior residence in high tuberculosis-incidence settings. This context provided a rare opportunity to examine the long-term immunologic associations of BCG in a contemporary, non-tuberculosis-endemic population.

PFAPA is widely considered an autoinflammatory disorder driven by dysregulated innate immune activation rather than persistent infection.[7–9] Consistent with prior reports, we observed a pronounced myeloid-skewed inflammatory profile preceding diagnosis, characterized by neutrophilia, elevated neutrophil-to-lymphocyte ratio, increased C-reactive protein and erythrocyte sedimentation rate, relative lymphopenia, and suppressed eosinophil counts.[7] These findings reinforce the concept that PFAPA represents a state of heightened innate immune responsiveness, potentially primed early in life and later manifested through recurrent febrile episodes.

The observed inverse association between BCG vaccination and PFAPA risk is biologically plausible in light of accumulating evidence that BCG induces durable reprogramming of innate immune cells through epigenetic and metabolic mechanisms, commonly referred to as trained immunity.[10,11] Experimental and human studies have shown that BCG exposure can recalibrate myeloid cell function, modulate inflammasome activity, and alter IL-1 family cytokine signalling pathways that are central to PFAPA pathophysiology.[12,15] In this framework, BCG may attenuate excessive innate immune reactivity or promote a more regulated inflammatory set point, thereby reducing susceptibility to autoinflammatory flares during childhood.

An additional post-hoc exploratory observation relates to the timing of BCG administration. Among the very small number of PFAPA cases with documented BCG vaccination, all vaccinations occurred within the first two days of life, whereas most vaccinated controls received BCG later in the neonatal period. Although based on extremely limited numbers and not suitable for formal inference, this pattern raises the possibility that the immunologic effects of BCG may be timing-dependent. Developmental studies suggest that immune imprinting and trained immunity responses differ substantially between the immediate perinatal period and later infancy.[10,16] Whether timing of BCG administration modifies long-term immune programming warrants further investigation.

Beyond BCG, additional vaccines showed associations with PFAPA risk in adjusted analyses, including modest inverse associations for MMR, MMRV and one DTaP–IPV formulation, and positive associations for rotavirus and bivalent oral polio vaccines. However, these estimates were heterogeneous, did not remain significant after false discovery rate correction and are likely to reflect residual confounding or chance rather than consistent vaccine-specific effects; they should not be interpreted as evidence to change existing immunisation practices.

This study has several strengths. The use of a nationwide healthcare database enabled inclusion of a large, well-characterized paediatric population with long-term follow-up. The availability of a dedicated, physician-selectable PFAPA diagnosis code within the LHS electronic health record allowed structured identification of clinically diagnosed PFAPA cases at the time of disease presentation and enabled assembly of a large, nationwide PFAPA cohort based on routine clinical practice. Requiring documentation of PFAPA as a chronic diagnosis that was not subsequently erased further enhanced diagnostic specificity. Rigorous matching of controls on age, sex, ethnic sector, socioeconomic status, and calendar time ensured comparable exposure opportunity and minimized temporal bias, as illustrated in the cohort construction flowchart. Vaccination data were extracted from a centralized immunization registry with high completeness, reducing misclassification of early-life exposures. The matched design, combined with conditional logistic regression and residual covariate adjustment, further strengthened internal validity.

Several limitations merit consideration. As an observational study, causal inference is inherently limited, and unmeasured confounding cannot be fully excluded. Factors related to early-life environment, household composition, or migration history may influence both BCG exposure and PFAPA risk. PFAPA case identification relied on clinical coding rather than standardized research criteria, an approach commonly used in healthcare databases that lack a unique identifier for this condition. However, the use of a dedicated, physician-selectable PFAPA diagnosis code within the LHS electronic health record supported structured identification of clinically diagnosed PFAPA cases, as reflected by the characteristic inflammatory phenotype observed.

The protective association observed for BCG vaccination was based on a small number of vaccinated PFAPA cases (3; 0.18%), whereas vaccination coverage was higher in the matched control group (398; 1.2%). In a matched case-control design, such marked differences in exposure prevalence between cases and controls are consistent with a strong potential protective association and do not indicate model instability when appropriate matched regression methods are applied; the findings are consistent with a lower occurrence of PFAPA among the vast majority of vaccinated children. Additional exploratory analyses of BCG vaccination timing, which were based on the small number of vaccinated cases, were conducted solely for hypothesis generation.

Despite these limitations, the consistency, magnitude, and biologic plausibility of the BCG association support the hypothesis that immune programming influences susceptibility to childhood autoinflammatory disease. These findings extend the growing literature on non-specific, long-term immunologic effects of BCG beyond infectious outcomes into the domain of paediatric autoinflammation. If replicated in independent datasets, these findings suggest that early-life immune programming may influence the risk of autoinflammatory fever syndromes, with implications for paediatric infectious disease practice and for the design of future vaccine or immunomodulatory interventions.

In conclusion, leveraging a unique national EHR resource with detailed historical vaccination data, we identified a strong inverse association between prior BCG vaccination and PFAPA risk. Although causality cannot be established, these findings provide epidemiologic support for a role of trained immunity and early immune programming in PFAPA pathogenesis and raise the possibility that immune modulation may represent a future avenue for mechanistic and translational research in this autoinflammatory disorder.

## Transparency declaration

### Access to data

Ariel Israel had full access to all the data in the study and takes responsibility for the integrity of the data and the accuracy of the analysis.

## Author contributions

Conceptualization: AI, EMa, YB

Methodology: AI

Investigation: AI, EMa, YB

Writing - original draft: AI, YB

Writing - review & editing: AI, YB, IG, AGC, SA, SV, EMa, EMe

## Declaration of generative AI and AI-assisted technologies in the manuscript preparation process

During the preparation of this work the authors used ChatGPT (version 5, OpenAI) in order to assist with language refinement for improved clarity and flow. After using this tool/service, the authors reviewed and edited the content as needed and take full responsibility for the content of the publication.

## Data Availability Statement

The raw (unaggregated) data supporting the findings of this study contain sensitive personal health information and are therefore not publicly available. Access to individual-level data is restricted by the institutional review board (IRB) and is permitted only to qualified researchers who receive IRB approval, in accordance with applicable regulatory and privacy restrictions.

